# Oncologists’ Experiences with Patient Death - a Qualitative Interview Study

**DOI:** 10.64898/2025.12.02.25341298

**Authors:** Minou Gandras, Maike Michalski, Svenja Wandke, Mareike Rutenkröger, Isabelle Scholl

**Affiliations:** Department of Medical Psychology, University Medical Center Hamburg-Eppendorf, Martinistraße 52, 20246 Hamburg, Germany; II. Department of Medicine, University Medical Center Hamburg-Eppendorf, Martinistraße 52, 20246 Hamburg, Germany

**Keywords:** Cancer care, oncology, end-of-life care, patient death, physician experiences, professional grief, coping strategies, medical education, workplace support

## Abstract

**Importance:** Frequent exposure to patient death may challenge oncologists’ well-being and compromise care quality. Despite these risks, evidence on how patient death affects oncologists across different healthcare settings remains limited.

**Objective:** To explore how patient death affects oncologists, focusing on personal and professional impact, coping strategies, and perceived support and educational needs.

**Design:** Qualitative study based on semi-structured telephone interviews conducted in 2024, analyzed using deductive-inductive content analysis.

**Setting:** Interviews with oncologists from various specialties, working in inpatient and outpatient adult cancer care settings across Germany.

**Participants:** Oncologists working within the German health care system with experience of at least one patient death were eligible to participate. Participants were recruited using purposive maximum-variation sampling. None withdrew after enrollment. The sample included a range of clinical roles and levels of experience.

**Intervention(s) or Exposure(s):** Cross-sectional observational study without intervention.

**Main Outcome(s) and Measure(s):** Participants’ descriptions of personal and professional effects of patient death, their coping strategies, and their reported support and educational needs.

**Results:** The sample included 23 oncologists (13 male) from diverse oncological specialties. Participants reported a wide range of personal and professional responses to patient death, from no impact to significant distress, but also described experiences of personal and professional growth. Emotional responses included sadness and helplessness, but also feelings of relief and satisfaction. Lasting effects, including shifts in clinical approach and life perspective but also adverse outcomes such as difficulty separating work from private life were reported. Common coping strategies included peer support, clinical reflection, professional distancing, and both team-based and individual rituals. Barriers to coping included taboos around death, blame dynamics, time constraints and unclear access to institutional support. While many reported no current unmet needs, the findings nonetheless revealed gaps in medical education and a demand for low-threshold support formats.

**Conclusions and Relevance:** Patient death can have lasting personal and professional impact on oncologists. Interventions addressing educational gaps and providing low-threshold support may help strengthen oncologists’ well-being and sustain care quality.

**Trial Registration:** Not applicable (qualitative observational study).

**Key points:** *Question:* How do oncologists experience and cope with patient death, and what support needs do they perceive?

*Findings:* In this qualitative interview study, oncologists described a wide range of personal and professional effects of patient death. Coping strategies included informal peer support, clinical reflection, professional distancing, and both team-based and individual rituals. While many participants reported no unmet support needs, the findings nonetheless revealed gaps in medical education and a demand for low-threshold support formats.

*Meaning:* Patient death can have lasting personal and professional impacts on oncologists, underscoring the need for accessible and structured support.

## Introduction

Cancer remains the second leading cause of death globally and the number of cases continues to rise^1^. Consequently, health care professionals (HCPs) in cancer care often face patient death. Therapeutic advances have prolonged treatment trajectories in many cancer types, including metastatic disease^2^, during which oncologists may develop emotionally close relationships with patients^3^. The death of a person with whom one has shared a close relationship typically elicits strong emotional responses^4^. Existing research suggests that physicians may be profoundly affected by patient death^5,6^ with some evidence indicating that such responses often remain unacknowledged due to prevailing social norms among physicians discouraging emotional disclosure^7,8^. Moreover, HCPs’ reactions to patient death appear to be characterized by some distinct features such as feelings of self-doubt, guilt, and failure^5,9^. Some authors have reported potentially beneficial effects of chronic exposure to patient death among oncologists, including clarifying life perspectives or increasing motivation to improve patient care^10^. However, repeated confrontation with patient death has also been associated with adverse mental health outcomes among physicians, including heightened distress, elevated rates of psychiatric morbidity^11^, burnout^12^, and substance abuse^13^. Compassion fatigue^13^ and secondary traumatic stress have also been reported among oncologists and can have negative consequences such as loss of empathy and emotional exhaustion^14^. Beyond their effects on physicians’ well-being, these psychological burdens may impair the quality of clinical care, including impaired physician-patient interaction^13^ or even withdrawal from patients near death^10^. Despite these known risks, only limited attention has been paid to oncologists’ experiences, their coping support needs, and perceived educational preparedness regarding patient death^15^. Moreover, existing studies on the impact of exposure to patient death on HCPs have largely been conducted in non-European contexts, with little data available from German healthcare contexts^16^.

Our study aimed to address this research gap by exploring the personal and professional impact of patient death on oncologists in Germany, their coping strategies, and unmet support needs. By doing so, we hope to shed light on the specific challenges and impacts faced by this professional group, identify areas of unmet support needs, and determine potential directions for future interventions.

## Methods

### Study design

A qualitative study using one-time, semi-structured telephone interviews was conducted, guided by the Consolidated Criteria for Reporting Qualitative Research (COREQ)^17^, see Supplement I.

### Sampling and recruitment

To ensure a diverse and relevant sample, participants were recruited through multiple channels. We emailed German Comprehensive Cancer Centers (CCCs), affiliated outpatient practices, and heads of clinical departments involved in cancer care at the University Medical Center Hamburg-Eppendorf (UKE). Department heads were asked to forward the invitation to their staff. The study was also promoted through social media posts by IS (senior author). Interested oncologists received further information from MG (first author) and were scheduled for an interview after providing informed consent. No participants withdrew after scheduling. Eligible participants were oncologists working in German adult cancer care who had experienced the death of at least one patient throughout their career.

A target sample size of n=20-25 was determined with the aim of achieving pragmatic saturation^18^, guided by the research group’s prior methodological experience, with recruitment concluding after 23 interviews. We used purposive maximum-variation sampling to capture a broad spectrum of oncologists’ perspectives, considering medical specialty, experience level, clinical roles, and care settings (i.e., inpatient and outpatient).

### Procedures

The study was approved by the local Ethics Committee (LPEK-0700). Written informed consent was obtained from each participant prior to data collection. Demographic data were collected through questionnaires and analyzed using SPSS. All interviews were conducted by MG, who had no prior close relationships with participants, from a private research office at UKE between April 26, 2024, and June 14, 2024. All interviews were audio-recorded (mean duration: 20.4 min; 11-33). A pilot interview with an oncologist, conducted to test the technical setup and the clarity of the interview guide, preceded data collection and was excluded from analysis.

### Interviews

A semi-structured interview guide (see Supplement II) was developed based on prior research on HCPs’ experiences with patient deaths^16,19^. Interviews began with an open question inviting participants to recall and describe a cancer patient’s death, followed by questions on general experiences, coping strategies, and unmet support needs.

### Transcription and data analysis

Audio recordings were transcribed verbatim in two steps. First, an automated transcription was generated using WhisperSpeaker^20^, a large language model-based tool combining open-source libraries such as OpenAI’s Whisper for speech recognition and pyannote.audio for speaker diarization. The resulting transcripts were then manually reviewed and corrected by MG and MM (co-author). Transcripts were pseudonymized using interview IDs that linked to demographic data (e.g., clinical role), with direct identifiers such as names and workplaces removed. Final transcripts were imported into MaxQDA^21^ for analysis.

Data were analyzed using a deductive-inductive coding approach with a preexisting coding structure from a related interview study by our research group^19^ with inductive adaptations, following a consented analysis plan (see Supplement III). MG and SW independently coded the first interview; MG, SW, and IS then reviewed the code structure. MG coded the remaining interviews and SW and MM revised the codes. Discrepancies were resolved through discussions. The full research team (MG, MM, SW, MR, IS) reviewed the evolving coding system at regular intervals. Participants did not receive transcripts or results for review.

## Results

### Sample Characteristics

All n=23 participants provided demographic data (see Table 1). The sample was gender-balanced (n=13 males, n=10 females) and represented diverse medical disciplines, clinical roles and experience levels. Most participants worked at university hospitals (n=22). Participants reported monthly averages of 60.6 treated (5-300) and 5.7 deceased (0.5-33) cancer patients. A mean of 27.8% (0-100%) of these deaths was perceived as distressing.

**Table 1.** Sample characteristics

|  | Participants (N=23) |
| --- | --- |
| *Characteristics* | *n (%)* |
| **Age group** |  |
| < 30 | 4 (17.4) |
| 31-40 | 14 (60.9) |
| 41-50 | 3 (13.0) |
| 51-60 | 1 (4.3) |
| > 60 | 1 (4.3) |
| **Gender** |  |
| Female | 10 (43.5) |
| Male | 13 (56.5) |
| **Work Setting** |  |
| University hospital | 22 (95.7) |
| Private hospital | 1 (4.3) |
| **Clinical setting<sup>a</sup>** |  |
| Inpatient services | 20 (58.8) |
| Outpatient services | 14 (41.2) |
| **Medical discipline<sup>a</sup>** |  |
| Medical Oncology | 13 (43.3) |
| Palliative Medicine | 5 (16.7) |
| Surgery | 3 (10) |
| Dermatology | 2 (6.7) |
| Gynecology | 2 (6.7) |
| Anesthesiology / Intensive Care | 2 (6.7) |
| Radiation Oncology | 1 (3.3) |
| Other | 2 (6.7) |
| **Physician role** |  |
| Resident physician | 10 (43.5) |
| Specialist | 7 (30.4) |
| Senior / Chief physician | 6 (26.1) |
| **Working experience (in years)** |  |
| < 5 | 11 (47.8) |
| 5-10 | 7 (30.4) |
| 11-20 | 3 (13) |
| > 20 | 2 (8.7) |

<sup>a</sup>Multiple selections were possible.

### Themes and Subthemes

In line with the aims of this study, the results focus on three main themes: 1) the impact of patient death on oncologists, 2) coping strategies, and 3) support needs. Tables 2-4 present identified themes, subthemes and illustrative quotes.

#### Theme 1: Impact of Patient Death on Oncologists

All participants recalled at least one memorable patient death when prompted. While most described some level of emotional response or distress, the extent and nature of impacts varied depending on several factors, with some participants reporting no relevant personal effect. Reported impacts were grouped into emotional and cognitive experiences, professional, and personal impacts (see Figure 1).

**Figure 1.**
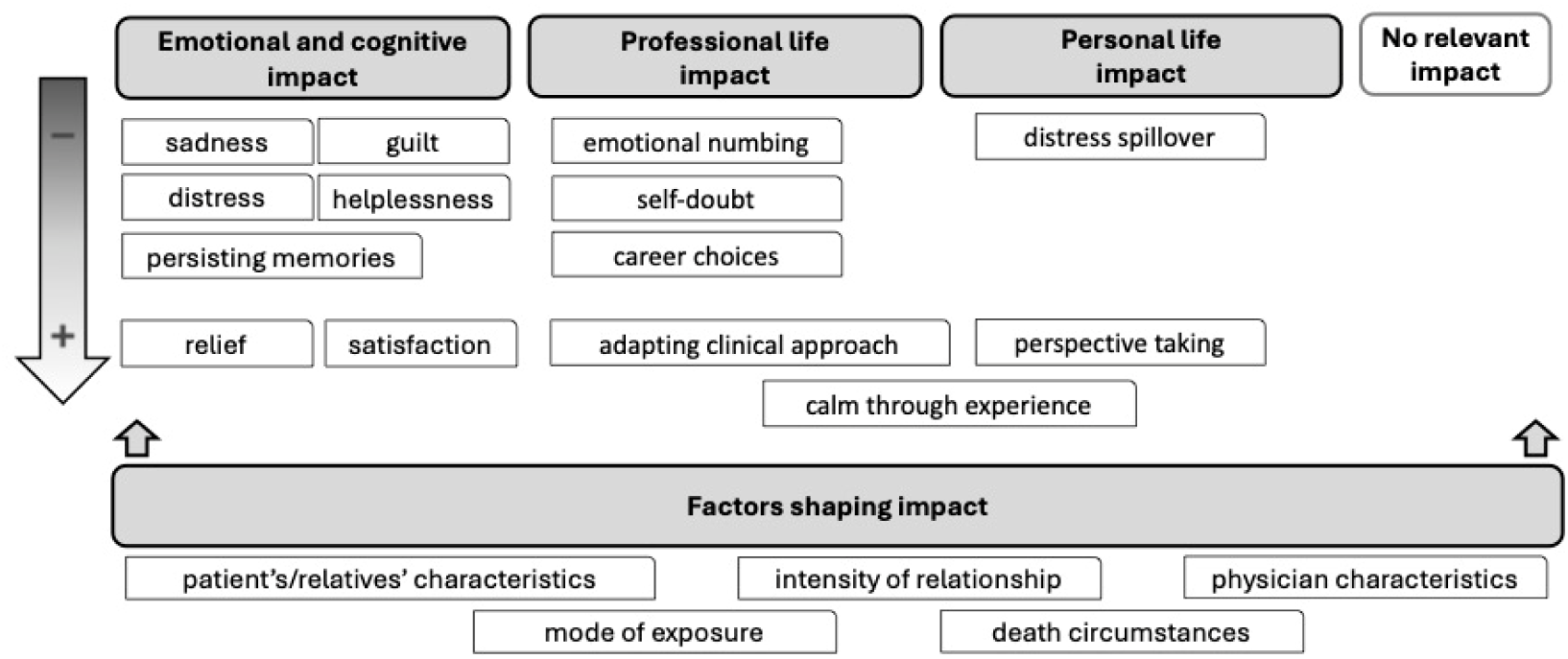
Impacts of experiencing patient death as reported by oncologists.

##### Emotional and cognitive impact

Participants reported varied emotional and cognitive responses to patient death. Sadness was described by most participants, while an experience of grief was described by some, typically characterized as less intense compared to private losses. Compassion with patients and relatives was common, with some oncologists expressing intensified fear of their own or loved ones’ death. Some, especially resident physicians, reported feelings of shock and helplessness. Guilt, anger or existential sorrow (*Weltschmerz*) were experienced by some, especially in response to young patients’ deaths. Positive emotions were also common, e.g., relief, satisfaction, and gratitude, especially after prolonged disease. Cognitively, many participants experienced recurring thoughts or vivid memories of deceased patients, sometimes persisting over years. After witnessing patient death, many participants felt more responsible and more conscious of life’s finality.

##### Professional impact

A common effect on professional functioning was adopting a calmer, more routine approach to patient death, often developing gradually over time and viewed as beneficial for patient care. However, few participants reported that it also carried negative aspects of emotional numbing and reduced empathy toward patients. Many participants reported self-doubts about their competence following patient death. Several participants described adapting their professional approach in response to patient death. This included shifting their idea of a “good death” - from preserving life at all costs to avoiding prolonged suffering and supporting patient autonomy in terminal stages - as well as initiating earlier conversations about death with patients and relatives. For some oncologists, patient death experiences shaped career choices: Five reported pursuing additional qualifications in palliative medicine and one partially withdrew from clinical patient care due to emotional distress.

##### Impact on personal life

Most participants reported positive effects on their personal lives, describing a perspective shift involving clarification of values, life priorities, and appreciation for their own lives. This was often attributed to increased awareness of life’s vulnerability following encounters with patient death. Several oncologists described feeling calmer and more prepared when facing death privately, with some engaging more proactively with death-related topics, e.g., discussing advance directives with relatives. Besides such positive aspects, however, difficulties separating professional and private life, i.e., “taking home” patient death-related distress, emerged. This theme was particularly salient among resident physicians.

##### Factors influencing impact

Participants identified factors influencing the level of distress following patient death, notably patient characteristics, such as young age, having young children, or difficulty accepting death. A close or longstanding physician-patient relationship also increased emotional impact. Factors related to patients’ relatives – such as familiarity with them, their distress or denial, or the absence of close relatives – were seen as increasing distress. The mode of death also emerged as a significant factor: Sudden death, rapid deterioration, failed resuscitation, poor symptom control (e.g., pain, dyspnea), dying alone, and certain treatment settings (e.g., intensive care units), were all linked to high distress. Physician-specific factors, including little clinical experience and similarity to patients (e.g., age, life circumstances), also emerged as contributing factors. Some oncologists mentioned systemic constraints limiting their ability to provide morally appropriate end-of-life care, especially in cases of prolonged aggressive treatment and unclear communication about a poor prognosis. These issues were often linked to hierarchical barriers that prevented oncologists from challenging senior colleagues’ decisions. Moreover, confrontation with limits of modern medicine added to distress, evoking feelings of powerlessness. Lastly, the mode of exposure to a patient death was considered significant with direct presence described as more emotional than retrospective notification.

#### Theme 2: Oncologists Coping with Patient Death

##### Interpersonal strategies

The most frequently identified resource of coping support was informal interaction with colleagues across different health care professions, sometimes involving the expression of emotions. Seeking social support in one’s private environment was less commonly reported. Some oncologists mentioned the availability of, and in fewer cases their own participation in, institutional support formats, including supervision and case discussion groups, and individual sessions with psycho-oncologists or chaplains. Additional interpersonal strategies included saying goodbye to the deceased patient’s body and contacting their relatives to offer condolences or express availability for further inquiries.

##### Intrapersonal strategies

Participants commonly reported reflecting on the medical case and their clinical performance and clarifying information gaps about death circumstances when necessary. Other salient strategies involved staying active, e.g., by returning to work quickly. Beyond behavioral coping forms, many oncologists reported adopting inward approaches, like accepting death and their emotional vulnerability. Another frequent subtheme was balancing emotional involvement and professional distance, e.g., by seeking distraction from clinical work, setting boundaries between work and personal life, or using humor to cope.

##### Rituals

Multiple oncologists described coping rituals performed alone or with colleagues. Individual rituals included prayers, closing patients’ eyes after death, and silently honoring the deceased by wishing them a good journey or recalling memories. Described team rituals ranged from informal acts, like searching obituaries together or placing symbolic lights outside rooms, to structured events such as team reflections or institutional memorial services. Those institutional rituals were mostly reported by palliative care physicians and were generally perceived as meaningful.

##### Coping Barriers

Participants identified several hindering factors to coping with patient death. These included limited chances to express emotions, a death taboo among physicians, and doubts about the legitimacy of their feelings, sometimes causing fear of showing vulnerability to colleagues or superiors and emotional suppression. Some noted a lack of institutional support or unclear access to it, while others reported low use of available resources, often due to time constraints. Another recurring coping barrier were unhelpful social interactions which hindered coping, professionally, due to lack of emotional engagement, focus on medical details, or dismissive responses; privately, due to others’ lack of understanding or emotional overwhelm. Few participants highlighted blame within teams after patient deaths and fear of legal consequences as additional barriers.

#### Theme 3: Oncologists’ Support and Educational Needs

Many oncologists reported no current unmet coping support needs. Nevertheless, some interviews revealed relevant gaps regarding educational preparation and support needs alongside clinical work. About half of the participants recalled some preparation for approaching patient death during medical school, mostly in early undergraduate stages as seminars on communication skills and delivering bad news. While some participants found these sessions helpful, others considered them too abstract. This was echoed in a need for more practical guidance during medical training, expressed by some oncologists. During postgraduate clinical training, most participants reported receiving no structured training in coping with patient death. Where such input did occur, it was often described as helpful, yet incidental – typically depending on senior physicians acting as role models or proactively addressing death-related issues. Many participants called for mandatory curricular content on patient death throughout medical education and expressed an interest in continuing education alongside clinical work, e.g., on end-of-life communication.

Besides educational aspects, participants also raised other current support needs in clinical practice. In hindsight, many wished for dedicated spaces at work for sharing experiences around patient death, even those who had initially declared no current unmet needs. Some called for team-based formats, including case discussions, supervision, or team rituals. Several also wished for a more open communication culture, particularly around uncertainty and mistakes. Lastly, individual counselling was requested by some, especially low-threshold, timely support offers, e.g., anonymous hotlines or drop-in sessions.

## Discussion

Our study illustrates the diverse effects of patient death on oncologists. While not all patient deaths were perceived as distressing, several factors shaped individual responses, including patient and physician characteristics, death circumstances, and the depth of the physician-patient relationship. Emotional responses ranged from grief and sadness, helplessness and guilt to relief and satisfaction. Long-term personal and professional consequences included calmness in facing death, changes of life perspective and clinical approach, but also difficulty maintaining professional boundaries and emotional numbing. Informal peer support and intrapersonal strategies, commonly involving medical reflection and emotional distancing emerged as central coping forms. Institutional resources, despite expressed interest, were rarely used – possibly due to reported barriers such as time pressure and taboos around death. While many oncologists reported no current support needs, educational gaps and a lack of accessible support formats emerged in some interviews.

In line with earlier non-European studies, participants experienced typical aspects of grief such as sadness, but also distinct HCP-specific aspects including professional self-doubt, guilt and fear of legal consequences following patient death^6,16,22^. The impact on oncologists appeared to be shaped by individual factors, rather than by the sheer number of patient deaths encountered, consistent with prior research^3^. While some oncologists found routine through exposure with death beneficial, others reported emotional numbing and reduced empathy, possibly indicating compassion fatigue among oncologists, a state of emotional exhaustion and reduced empathy resulting from prolonged exposure to others’ suffering^23^, in line with findings from other healthcare systems^3,13^. Several participants described focusing on medical aspects during case reflection and quickly returning to work tasks as a coping strategy. This has been discussed as a form of emotional compartmentalization which – while potentially necessary for short-term professional functioning – may compromise physician wellbeing in the long term^8^.

Although many participants felt adequately supported in coping with patient death, our results highlight specific gaps in educational and institutional support, as well as systemic barriers to participation. Reported unmet needs underline the importance of systemically integrating practice-oriented curricular contents on patient death during medical education and clinical training, which – consistent with medical education research – should be adapted to learners’ experience level and supported by pre-and debriefing to ensure psychological safety^24^. Some adverse impacts, notably feelings of helplessness and difficulty maintaining professional boundaries, appeared particularly common among resident physicians. Consistent with prior research^14^, early-career oncologists may be especially vulnerable to emotional overwhelm following patient death and may require specifically tailored support interventions to prevent adverse outcomes or maladaptive coping in the long term. Although interest in hospital-based coping support was widely expressed, low uptake of such offers was reported. This apparent discrepancy may stem from coping barriers described by participants, including time constraints, perceived taboos around death, blame culture, and reluctance to display vulnerability around colleagues and superiors. Interventions targeting communication culture around death and error could improve both coping and acceptance of available offers. Moreover, institutions may consider the possibility of establishing low-threshold, timely individual counselling formats such as open consultation hours or hotlines. Informal peer support and emotional expression emerged as key coping resources, though availability varied, with some oncologists describing social interactions as unhelpful. Moderated or regular peer group formats may enhance consistency and reduce reliance on incidental support.

### Strengths and limitations

To our knowledge, this is the first study exploring oncologists’ experiences with patient death in the German healthcare system. Our sample was gender-balanced and included diverse medical specialties, clinical roles, and experience levels. Data analysis followed a pre-agreed protocol, was conducted by three researchers, and included regular quality reviews within the full research team. We adhered to the COREQ reporting standards.

Limitations include a sample predominantly consisting of oncologists from university hospitals, with only one from a private hospital and none from outpatient practices or faith-based hospitals. Thus, despite purposive maximum-variation sampling, some oncologist perspectives may remain underrepresented in this study. Furthermore, self-selection bias may have occurred due to convenience sampling. Consequently, while not the primary objective of qualitative research, generalizability is limited by these factors.

## Conclusions

This study provides new insights into the varied extent and nature of patient death’s effects on oncologists, often impacting both personal and professional domains. While distress was a common effect of being confronted with patient death on oncologists, it also prompted reflection, personal and professional growth. Informal peer support, preserving own professional boundaries, and clinical reflection emerged as valuable coping strategies, yet gaps in medical education and institutional support became apparent – particularly a lack of practice-oriented training for students and professionals and easily accessible coping support formats. These findings offer a foundation for further research to quantify unmet support needs, evaluate preferred support frameworks, and develop strategies to detect and support oncologists at risk of adverse outcomes.

## Article information

### Author Contributions

IS and MR are the responsible principal investigators of the study and share senior authorship. IS, MG, MM, SW and MR were involved in the planning and preparation of the study. MG recruited participants and collected data, and analyzed the data with the help of MM and SW. All authors interpreted the results. MG wrote the first draft of the manuscript. MM, SW, MR and IS critically revised for important intellectual content. All authors gave final approval of the version to be published and agreed to be accountable for the work.

## Supporting information

Supplement IV: Tables_2-4_Example quotations

Supplement I: COREQ Checklist

Supplement II: Interview Guide

Supplement III: Coding Tree

## Data Availability

All data produced in the present study are available upon reasonable request to the authors

