## Supplement IV: Tables_2-4_Example quotations for "Oncologists’ Experiences with Patient Death - a Qualitative Interview Study"

Table 2.

Theme 1: Impact of patient death on oncologists

| **Subthemes** |  | **Example quotations** |
| --- | --- | --- |
| Few/no effects |  | „I don’t feel burdened by it [patient deaths] in a negative way.” (ID02, specialist in medical oncology and palliative medicine  “I never felt deep regret that these patients had died, because I’m simply very much in my professional role.” (ID11, senior/chief physician in radiation oncology) |
| Being touched |  | “That [patient death] doesn’t leave me untouched, even in my professional role.” (ID01, senior/chief physician in medical oncology) |
| Being distressed |  | “It’s pretty tough, watching people die, I have to say. That makes this profession very, very hard.” (ID14, specialist in surgery)  “Still, there are always patients whose deaths burden you emotionally.” (ID15, resident physician in medical oncology) |
| Doubts about the legitimacy of own emotions |  | “I guess you just have to accept it somehow because after all, you don’t want to burden others with all your stories.” (ID05, resident physician in medical oncology) |
| **Emotional impact** |  |  |
| Satisfaction |  | “But during the dying process, we can alleviate suffering and prolonged distress. And if we’re able to do that, then from a physician’s perspective, it really is a meaningful and positive experience to be able to provide that kind of support.” (ID06, resident physician in anesthesiology/intensive care) |
| Relief |  | “I think what is actually predominant is a sense of release and relief, because it usually happens over a long period of time and is not a sudden death.” (ID23, resident physician in medical oncology and surgery) |
| Gratitude/appreciation |  | “Sometimes I actually find remembering [a deceased patient] to be something beautiful because it means they’re not forgotten. This way, you preserve the memory of someone you cared about.” (ID14, specialist in surgery) |
| Existential sorrow (*Weltschmerz)* |  | “[...] and also just a feeling of unfairness, why it had to happen to such a young person in particular.” (ID12, resident physician in medical oncology) |
| Overwhelm/helplessness |  | “And when [the patients] expressed fear or something like that, I sometimes felt overwhelmed, being there and not really able to ease their fear.” (ID20, resident physician in dermatology) |
| Sadness |  | “I do always find it sad and depressing. And I have to say honestly, even though I’ve been doing this for a very long time and see many [...] dying patients, when it gets to that point, it still often tightens my throat, without wanting to sound melodramatic.” (ID08, senior/chief physician in gynecology) |
| Shock |  | “Well, in her case it was actually foreseeable, but still, when you get the news -- her husband had called -- at that moment, I was shocked.” (ID20, resident physician in dermatology) |
| Anger |  | “Beyond that, I also had feelings of anger because the patient died during the night on an intensive care unit, where the physician on duty, of course, wasn’t aware of the significance, or the full scope, so to say, of the patient’s journey.” (ID05, resident physician in medical oncology) |
| Guilt |  | “I think it’s a very dangerous thing when, as a physician, you start asking yourself questions of guilt. Because I believe that can be very distressing.”  (ID23, resident physician in medical oncology and surgery) |
| Grief |  | “And, well, to put it quite simply, grief is always there [when a patient dies]” (ID12, resident physician in medical oncology) |
| Compassion |  |  |
|  | For patients | “I don’t say find it distressing in the sense that I take it home or feel weighed down myself, but I find it so sad for the patients.” (ID02, specialist in medical oncology and palliative medicine) |
|  | For relatives | “And what makes me feel the most sorry and uneasy is when I speak with the relatives afterwards.” (ID08, senior/chief physician in gynecology) |
| Fear of one's own finiteness |  | “Over the years, you naturally accumulate more fears, you have to say. Because when you get to know young patients with serious illnesses [...], of course, you also start thinking about what that would be like for yourself, for your family. That is something that tends to increase.” (ID14, specialist in surgery) |
| **Cognitive impact** |  |  |
| Long-term vivid memories of deceased patients |  | “The case that immediately comes to mind is one of end-of-life care from my first year of professional practice, which has shaped my entire professional life and career path.” (ID22, senior/chief physician in medical oncology and palliative medicine) |
| Recurring thoughts of deceased patients |  | “There are, of course, as I said, patients [...] you keep thinking about again and again later on [after their death].” (ID14, specialist in surgery) |
| Sense of responsibility |  | “I am very aware that you only die once and that you don’t get a chance to make up for what you -- as the treating physician -- missed, if you don’t support, ease, and accompany someone’s dying optimally.” (ID01, senior/chief physician in medical oncology)  “That [a patient death] is distressing, yes, but what’s even more distressing is when we’ve done something wrong.” (ID06, resident physician in anesthesiology/intensive care) |
| Conscious experience of life’s finality |  | “When life actually leaves the body, there always is a certain feeling, how should I put it... when that finality becomes clear, it's still a very, very special feeling. That doesn’t go away, even after accompanying many deaths [...] -- this awareness of finality has something to it, because it really is one of the very few things that cannot be undone.” (ID07, senior/chief physician in medical oncology and palliative medicine) |
| **Professional impact** |  |  |
| Routine in dealing with the patient death |  | “You just kind of get used to how it [patients dying] works, and I think that is an internal automatic process.” (ID05, resident physician in medical oncology)  “And of course you become more confident and, I think, less distracted by other things. Instead, you focus on what’s really important for the [dying] patient at that moment.” (ID01, senior/chief physician in medical oncology) |
| Proactive communication with patients about death and dying |  | “I do feel that, as a consequence, I realized how important this [death] is, and since then I address it early on, and not like ‘you’re going to die tomorrow’, but more generally: How would you imagine it? What could we do to create a sort of environment from the beginning?” (ID10, specialist in medical oncology) |
| Decision to pursue additional training/further education |  | “Professionally, I have to say that a very bad death in my old hospital [...] where I worked as a resident, actually led me to choose the specialty I did, namely palliative medicine.” (ID18, specialist in palliative medicine) |
| Reduction of work in clinical patient care |  | “So the impact [of patient deaths] is that thoughts have kept coming up about whether I should withdraw more from clinical and direct patient care.” (ID01, senior/chief physician in medical oncology) |
| Self-doubts about one’s competence |  | “Because it does sometimes make you think, like, wow, they all die under my care, I am the treating physician -- could I have done something better, could I have done something differently?” (ID05, resident physician in medical oncology) |
| Decreased emotional resonance/empathy towards patients |  | “But apart from that, it [a patient’s death], as harsh as it sounds, is acknowledged, filed away, checked off, sorted out.” (ID08, senior/chief physician in gynecology)  “Probably most of all numbness, that you don’t let it get to you emotionally as much anymore.” (ID09, resident physician in surgery). |
| **Personal life impacts** |  |  |
| Greater appreciation for one’s own life (perspective taking) |  | “Yes, I do think that I actually decided for myself that things shouldn’t be postponed so much anymore. Because this experience of sudden death [...] we see that again and again. That has changed my mindset a bit, in the sense of non-postponability of things -- that you shouldn’t wait until retirement to do things, and so on.” (ID07, senior/chief physician in medical oncology and palliative medicine)  “This deep, deep gratitude for the health of my family and friends, my child, my husband, my parents, and especially myself -- that I am constantly reminded through my daily work of what else can be. And that certain things then just stop being so important.” (ID18, specialist in palliative medicine) |
| Changes in beliefs related to death |  |  |
|  | Dissolution/ loss of previous beliefs | “In the past, I would have just said: okay, live as long as possible, no one wants to die. But now, when I care for patients, it’s not about postponing death anymore but more about the patient being able to live autonomously until the end, express their wishes, maybe die surrounded by family or at home.” (ID09, resident physician in surgery) |
|  | Consolidation/ clarification of one’s own beliefs | “That has perhaps become even stronger, that I find it is best for a patient to be awake, or at least more awake, and to consciously approach the process [of dying], rather than dying out of an anesthesia or a coma that has been ongoing for some time.” (ID06, resident physician in anesthesiology/intensive care) |
| Serenity in dealing with death and dying |  | “But it is definitely the case that what I’ve learned in the hospital has also taught me how to deal with dying in my private life, that I’m able to accompany people. That I’m not afraid to be there, that I’m also not afraid to be present when someone has died, so to speak.” (ID14, specialist in surgery) |
| Active engagement with death in one’s private life |  | “Yes, and through my work in oncology and my engagement with the topic [death], I have definitely tried to make sure that certain things are clarified in advance, for example, an advance directive, or that certain steps have already been taken.” (ID12, resident physician in medical oncology) |
| Difficulties in separating personal and professional life |  | “And that [a patient death], took a lot of strength out of me, and I carried a lot of it home with me, in the sense that I also grieved over it at home.” (ID05, resident physician in medical oncology) |
| **Distinction between experiences with death in professional and personal contexts** |  |  |
| Difference in the duration of emotional impact |  | “When it is someone close to you [from one’s personal life], it really can tear your heart out. And it takes much, much longer to process.” (ID23, resident physician in medical oncology and surgery) |
| Difference in intensity |  |  |
|  | Due to greater emotional regulation in response to patient death | “And in the professional setting, it [dealing with death] was much easier for me, because, even though there is also a taboo there, I was somehow better able to process it.” (ID10, specialist in medical oncology) |
|  | Due to less close personal connection with patients | “So, [there are] definitely differences -- with family members, of course, there is a completely different kind of emotional connection.” (ID20, resident physician in dermatology) |
|  | Professional role affected vs. entire self affected | “I think there is a very clear difference, because of course, you’re more affected by deaths in your private life. In that case, I am not professional, at work, I am professional.” (ID03, specialist in palliative medicine) |
| Commonality: Experience of grief/sadness |  | “The commonality [between patient deaths and deaths in personal life] is definitely always a feeling of grief.” (ID05, resident physician in medical oncology) |

| **Factors influencing perceived impact of patient deaths** |  |  |
| --- | --- | --- |
| Related to the mode of exposure to a patient death |  | “Often you just get a message saying something like, ‘By the way: patient X-Y passed away on such and such date’, and of course that’s a much more anonymous and far less intense experience than, for example, accompanying a patient on the ward, perhaps even providing end-of-life care. That’s a much more involved care relationship, and you experience completely different emotions.” (ID12, resident physician in medical oncology) |
| Confrontation with the limits of medicine |  | “And also, [I feel] a certain frustration with medicine not being advanced enough to help this person more with their illness. That’s something you experience a lot in oncology: At some point, you reach your limits and can no longer help the patient and you tell them the tumor has grown, and there’s nothing more we can do.”(ID05, resident physician in medical oncology)  “You believe you can somehow do everything, but in the end, reality shows that we simply can’t help many patients the way we imagined.” (ID12, resident physician in medical oncology) |
| Constraints due to systemic conditions |  | “You really have to fight to ensure that people can die the way they want to. That has changed. You have to challenge the system [to prevent] that the last catecholamines are pumped in and we keep it going for so long because none of us dares to stop it.” (ID21, specialist in medical oncology) |
| Patient care not possible according to own moral concepts |  | “And of course I’m not the boss, I’m not a senior physician, I’m not the one who has the final say. And from the more senior side, it was always: ‘Let’s still try this.’ It was a terrible situation for the patient, for us, for her husband. Because we all knew there was no hope.” (ID14, specialist in surgery) |
| Related to clinical setting or medical specialty |  | “We talk a lot and very openly about it [patient deaths]. But that’s different in the professional environment outside of palliative medicine.” (ID22, senior/chief physician in medical oncology and palliative medicine)  “I find that in oncology, everything is always very calm and it’s clearly a completely different setting. In intensive care units, it’s always a matter of all or nothing.” (ID21, specialist in hematology/oncology) |
| Related to the frequency of experienced patient deaths |  | “I think that dealing with death on a daily basis changes you differently than ­­-- so to say – experiencing one every quarter.” (ID04, specialist in gynecology) |
| Related to the nature of death/way of dying |  | “[The patient] had symptoms of severe fear of death and shortness of breath due to blood loss. The patient had frequent, heavy bleeding into the intestinal tract and developed anemia. Naturally, he felt absolutely awful and he looked like it. You could see it right away, he was completely pale. And of course, I felt deeply sorry for this patient.” (ID11, senior/chief physician in radiation oncology)  “Just a few months earlier, [the patient] had actually been doing really well. Then everything happened very quickly, from the first pain to the diagnosis. In the end, I think it was about four weeks until he died. He was a patient who was really in the middle of life.” (ID13, resident physician in dermatology) |
| Related to the physician-patient relationship |  | “Especially with patients you’ve cared for over a longer period, when they die, you do feel relatively close to them.” (ID23, resident physician in medical oncology and surgery) |
| Related to patient characteristics |  | “What I find more difficult is accompanying patients [...] who can’t let go and still have something in their lives left unfinished, and you simply can’t help them with that.” (ID02, specialist in medical oncology and palliative medicine)  “When it comes to younger patients, I have to say that dying [...] tends to be more distressing for everyone present, the relatives as well as the nursing and medical staff.” (ID16, resident physician in medical oncology) |
| Related to characteristics of the patients’ relatives |  | “There are so many people on the ward in their 70s or 80s who have no one -- and then it becomes incredibly difficult to experience a good death, because they simply don’t have anyone left. In the end, I believe that, as a human being, it comes down to not feeling lonely.” (ID10, specialist in medical oncology)  “There was also the fact that [the patient] had a partner [...] who was always very distressed. When he couldn’t come to the outpatient clinic because he was in the emergency room, she would sit with me, crying.” (ID12, resident physician in medical oncology) |
| Related to characteristics of the oncologist |  | “I could kind of identify with the woman, because she was in a similar situation to mine with her child. And therefore the case touched me.” (ID03, specialist in palliative medicine)  “But back then I was still quite young and, so to speak, a bit naive. And it really took a lot of strength out of me; I carried a lot of it home with me.” (ID05, resident physician in medical oncology) |

Table 3.

Theme 2: Oncologists’ coping strategies

| **Subthemes** |  | **Data extracts** |
| --- | --- | --- |
| **Interpersonal strategies** |  |  |
| Professional support |  |  |
| Participation in group/team formats |  | “And I must also say that I find ward conferences, where the different staff members from the various responsible areas come together to exchange, very helpful. I have always considered that helpful.” (ID19, resident physician in medical oncology) |
| Support from chaplains |  | “And for religious people, there are also chaplains, for example, we have a chaplain at our clinic.” (ID04, specialist in gynecology) |
| Support from psychologists |  | “In our clinic, we have psycho-oncologists, especially for the dying process, and we can also talk to them ourselves.” (ID20, resident physician in dermatology) |
| Informal social support |  |  |
| Support from colleagues |  |  |
|  | From physicians | “Yes, in the professional setting, that [peer exchange] is often something common, especially when we are treating the same patients. And in those cases, the focus is more on the clinical aspect, on trying to understand why someone died -- especially when things happened suddenly. It is usually approached on a more objective level.” (ID07, senior/chief physician in medical oncology and palliative medicine)  “Most of the time, we notice it in each other – for example, when someone comes back from a conversation with relatives or a patient, someone might ask: ‘Wow, what’s going on, are you okay? Was that tdifficult?’ Or you might say it yourself, like: ‘That was really hard.’” (ID18, specialist in palliative medicine) |
|  | From nursing staff | “That was when the tears really came, and then a nurse came over, who must have noticed it during handover, and said: ‘Would you like to talk about it?’ And then, somehow, something inside me broke open.” (ID02, specialist in medical oncology and palliative medicine) |
|  | From psychological colleagues | “I actually find it very interesting to talk about it again with independent colleagues, with psycho-oncologists or psychologists.” (ID21, specialist in hematology/oncology) |
|  | From other professional groups | “But since we are very closely connected on our wards -- psycho-oncologically, therapeutically, in nursing, and so on -- and we work in close interdisciplinary exchange, I am very lucky that we receive a great deal of support as a team when something becomes distressing.” (ID02, specialist in medical oncology and palliative medicine)  “I often do that too, that I, for example, frequently talk to our art therapist about cases.” (ID18, specialist in palliative medicine) |
| Support from private social circle |  | “And of course, my family is also a source of support. Especially my husband, because he also worked in the medical field and knows all of this. And then of course, when I bring patient cases home, he listens and gives me space to talk about it.” (ID14, specialist in surgery)  “I also talk a lot with my family about it who are -- I find -- always easy to talk to, even though none of them are doctors and they only know this from a personal perspective. But I think you really do get a lot of support just by talking to someone about it.” (ID20, resident physician in dermatology) |
| Expression of emotions |  | “Everyone is actually relieved when they can share it, when you do not have to carry it around all by yourself.” (ID14, specialist in surgery)  “Of course, without naming anyone, but very often it is simply a matter of having talked about it once. Like: ‘This is weighing on me, that was a difficult case, or it really affected me for such and such reasons.’ That is usually enough, and it works quite well.” (ID18, specialist in palliative medicine) |
| Saying goodbye to the body |  | “So, when I perform the post-mortem examination, I actually speak to the patient -- or to the body. Not extensively, but I do say goodbye, and especially if I knew them well, I thank them for having met them and wish them all the best, for example.” (ID18, specialist in palliative medicine) |
| Initiating contact with patients’ relatives |  | “Sometimes I offer the relatives to come see me again after a few days, if they feel the need to talk, so that we can just sit down together and talk again.” (ID08, senior/chief physician in gynecology)  “When I have accompanied patients for several years and seen the family again and again, then it does happen that we exchange condolences.” (ID15, resident physician in medical oncology) |
| **Intrapersonal strategies** |  |  |
| Activities |  |  |
| Professional reflection on patient death |  | “To what extent was I involved in the treatment? To what extent did something maybe go wrong medically? So I tend to think about it more, yes, a bit mechanically in professional terms.” (ID06, resident physician in anesthesiology/intensive care) |
| Closing information gaps about patient death |  | “I like to read up on it. If I wasn’t directly involved or had a day off and then hear about it the next day, I usually do go back and check again, read through how it happened, how it took place.” (ID03, specialist in palliative medicine) |
| Continuing work tasks |  | “[When I] find out that the patient has died, I pause for a moment, but then, especially when you find out during hospital work, you quickly return to routine. And it’s a bit different when you accompany the death directly, but even then, it is often the case that the next thing comes up right away and you move on to the next task.” (ID01, senior/chief physician in medical oncology) |
| Lighting a candle |  | “At the front desk of the ward, a small candle, an LED candle, of course, is lit for as long as the patients or the deceased remain on the ward.” (ID02, specialist in medical oncology and palliative medicine) |
| Visiting a spiritual space |  | “Yes, for me, as I am a religious person, it [a place of coping support] would be the church.” (ID03, specialist in palliative medicine) |
| Physical activity |  | “Since sports is a good way to balance things out, for me, it is sports [that helps cope with patient deaths].” (ID03, specialist in palliative medicine) |
| Adapting one’s attitude |  |  |
| Consciously allowing emotional connection and vulnerability with patients |  | “The moment where I stop doing that [bonding with patients], I might as well stop being a physician, because that is actually the reason why I became one. I bond with people as much as I want to, as long as I can somehow cope with it. But of course, when they die, it may be different than if one had no connection at all.” (ID05, resident physician in medical oncology) |
| Applying own beliefs about life and death |  | “I myself have a Christian background and I would say that it helps me. And I also see that it helps patients when they have a religious belief that offers some kind of answer to the question of what happens when I die.”  (ID11, senior/chief physician in radiation oncology)  “I want to believe that these [deceased] people are somehow still around somewhere, even if they only live in our hearts, that one just doesn’t completely disappear.” (ID14, specialist in surgery)  “I believe there is nothing after that, I don’t think you go to heaven or get reincarnated somehow. I believe there is nothing afterwards.” (ID23, resident physician in medical oncology and surgery) |
| Acceptance |  | “At some point you see: Patients dying is simply part of it. And it is not a sign of medical incompetence, but it is a natural process that, if done well, can be accompanied in the best possible way.” (ID16, resident physician in medical oncology) |
| Finding the right balance |  |  |
| Letting time pass |  | “Time heals all wounds.” (ID11, senior/chief physician in radiation oncology) |
| Pausing/taking a moment |  | “It is often the case [after a patient death] that I pause for a moment and think of the person.” (ID14, specialist in surgery) |
| Seeking distraction or balance |  | “Yes, following my hobbies, or, I don’t know, doing other things [helps]. I am a clinician scientist, and research is fun too and that is, so to speak, a world full of tubes and not people all the time […] and of course, sports, I don’t know, movies, friends, all these things.” (ID05, resident physician in medical oncology) |
| Seeking physical distance from work |  | “I only work part-time, so that I can also keep some distance from all of it.” (ID03, specialist in palliative medicine) |
| Maintaining a professional distance |  | “But as physicians, we are supposed to, or rather we have learned, not to suffer with the patients but only to feel with them.” (ID06, resident physician in anesthesiology/intensive care)  “I do try not to put myself too deeply into the patient’s position, like thinking: How would I feel if this were my father? Because that is the barrier I have built for myself, which would collapse otherwise.” (ID16, resident physician in medical oncology) |
| Clear separation between personal and professional life |  | “But I do not take it home with me. Once I finish work, I do not really think about it anymore or discuss it at home. It basically stays at work.” (ID12, resident physician in medical oncology) |
| Humor |  | “I would say, in fact, a trace of black humor [helps me cope].” (ID02, specialist in medical oncology and palliative medicine) |
| **Rituals** |  |  |
| Googling obituary notices |  | “I don’t really know why, but my colleagues and I always do that together, because we have cared for many [deceased patients] jointly, that we google the obituaries one more time.” (ID20, resident physician in dermatology) |
| Acknowledgement/honoring the deceased patient |  | “When I learn about it, I try to remember the patient and reflect on the time we shared, if one can put it like that. And I simply wish him or her peace inwardly, and their relatives too.” (ID23, resident physician in medical oncology and surgery) |
| Team-based/institutional rituals |  | “Yes, of course that is something very specific to our ward, that we have certain rituals. It is quite nice; we place lanterns in front of the doors of the rooms where the [deceased] patients are lying.” (ID02, specialist in medical oncology and palliative medicine)  “In our teams we also have a kind of remembrance once a week, where we name all those who have died, and our chaplain says something and expresses a wish.”(ID07, senior/chief physician in medical oncology and palliative medicine) |
| Praying |  | “Back when I was a young physician, I would sometimes say a short prayer.” (ID18, specialist in palliative medicine) |
| **Coping barriers** |  |  |
| Lack of professional support services |  | “There is no team-based processing. Of course, one can talk to colleagues, but there is no professional or structured review of certain situations.” (ID10, specialist in medical oncology)  “No, unfortunately, ultimately, the kind of supervision one might sometimes need does not exist.” (ID12, resident physician in medical oncology) |
| Taboo around death |  | “Sometimes people do not dare to talk about it -- that is also why you are doing these interviews -- the topic is very, very taboo, completely taboo, truly. I mean, the only ward where it is not taboo is the palliative care ward. That is why all the colleagues constantly prefer to request a palliative medicine consultation, just to avoid dealing with it themselves.” (ID10, specialist in medical oncology)  “It [patient death] is not a topic. No one has ever asked anything like ‘How are you doing with this?’ or ‘Is this somehow distressing for you?’ It is simply not addressed at work by superiors. Not at all.” (ID17, resident physician in medical oncology) |
| Limited acceptance of existing professional support services |  | “What I keep thinking about, or what I find quite alarming, is that in medical oncology […] -- It is not that Balint-groups do not exist, but I find it alarming how little they are used. And I myself am actually also among those who always prioritize all other things.” (ID01, senior/chief physician in medical oncology)  “I believe it [an institutional coping support offer] was somehow discontinued now due to a lack of people’s interest. I would wish that more people would participate, because it is actually a good offer. It is certainly helpful for everyone. But well, if no one shows up and participates, how should they keep offering it? That is of course difficult.” (ID05, resident physician in medical oncology) |
| Lack of awareness of support services and access pathways |  | “No, I have no idea. Probably, I could somehow ask around, through the colleagues on the palliative care unit, but I would not know where to turn right now, no.” (ID17, resident physician in medical oncology)  “There are probably some [institutional coping support offers], but I do not know them. But I would be interested to know who I could turn to in theory. I cannot imagine that there is nothing, but I am not aware of any concrete offers, and I find that unfortunate.” (ID23, resident physician in medical oncology and surgery) |
| Sharing experiences perceived as unhelpful |  |  |
| With people from private life |  | “In my private life, I have become reluctant to talk about it, because I feel that the benefit is almost zero -- when people respond with questions like ‘Oh God, that is terrible, how do you even manage to deal with that?’ -- for me, for processing it.” (ID01, senior/chief physician in medical oncology) |
| With colleagues |  | “There have been situations at the hospital, where, for example, I had a really difficult death during the night, and in the morning handover something like, ‘well, it was probably for the best,’ was just said, without addressing it in any way. It was treated as if someone had been discharged, and no one really addressed it and in those moments, I sometimes did not feel understood.” (ID20, resident physician in dermatology)  “In professional settings, things are sometimes handled a little --, I do not want to say carelessly, but for some reactions that might actually be the right word. When I think back to situations in the intensive care unit, there were sometimes reactions that felt dismissive. I find that very disturbing and lacking in respect.” (ID21, specialist in medical oncology) |
| In supervision |  | “I have to say that supervisions do not really help me with that [coping]. We do have regular team supervisions in a larger team […]. It is too far removed. It needs to be addressed directly, and then it is fine.” (ID18, specialist in palliative medicine) |
| No expression of emotions |  | “I do not do that [talk about patient deaths] in my private life, not at all with my partner, because she does not want that emotionally. Yes, it’s not possible. Most cases that truly distressed me or moved me, or that I continued to think about for a longer time, I rather dealt with on my own.” (ID16, resident physician in medical oncology)  “I have never talked to anyone about it [patient deaths].” (ID23, resident physician in medical oncology and surgery) |
| Time pressure |  | “I have also experienced situations where someone wanted to finish their handover quickly, and you could see on the monitor that someone was dying, right there in the intensive care unit, and then it sometimes seemed to have little importance.” (ID07, senior/chief physician in medical oncology and palliative medicine)  “I know that there used to be some offers where people met regularly at fixed times in larger groups, but that is honestly difficult to reconcile with clinical routine.” (ID12, resident physician in medical oncology) |
| Pressure to meet expectations/perfectionism in dealing with patient death |  | “You are somehow so sad -- but then it is like, ‘well, now it really has to move on, you are allowed to be sad for three days, but then it has to continue’.” (ID10, specialist in medical oncology) |
| Suppressing one’s own emotions |  | “What I have always found unhelpful is this ignoring and pushing away. It is not something anyone specifically said to me, but it is a tactic many of my colleagues use, to block out death.” (ID14, specialist in surgery) |
| Dishonest communication with patients about prognosis |  | “And also, to some extent, the way superiors handled things, giving patients false hope and continuing therapies that were, in truth, futile. That is actually the worst part of it all.” (ID13, resident physician in dermatology) |
| Blame dynamics |  | “And it is always just about ‘Who is to blame for that person’s death?’” (ID10, specialist in medical oncology)  ”Blaming each other among colleagues and not taking responsibility -- I find that to be a very poor style. That really, really bothered me.” (ID12, resident physician in medical oncology) |
| Perceived threat of legal consequences from treatment decisions |  | “I think that everyone is afraid of being questioned somehow. That is a bit what medicine today is about: The main thing is not to get sued.” (ID10, specialist in medical oncology) |

Table 4.

Theme 3: Oncologists’ support and educational needs

| **Subthemes** | **Data extracts** |
| --- | --- |
| “Were oncologists educationally prepared to cope with patient deaths?” |  |
| Yes,  during medical school |  |
| Pre-final year | “Certainly, there are seminars in medical school about delivering bad news and courses on palliative medicine, but you are only truly confronted with it once you begin working in the profession. At least for me, I had to engage with the topic on my own. On paper or in theory, it is one thing, but in practice it is something else.” (ID16, resident physician in medical oncology)  “I was among the first cohorts at my university to already have palliative care, but only as a small component in the tenth semester. […] Also all of those […] communication models, how to talk -- that helped me a lot. The lecture helped me a lot as well.” (ID18, specialist in palliative medicine) |
| Final clinical year | “In terms of the formalities, yes: How does the post-mortem examination work, what needs to be filled out? Emotionally, I would say no.” (ID11, senior/chief physician in radiation oncology) |
| Yes,  during postgraduate specialist training (during residency) | “Back then I had a very good mentor, the senior physician in the intensive care unit, who provided very patient-centered care to the patients and relatives. I observed this and experienced it because I was present during his conversations. But he also explained to me in advance exactly how he does it.” (ID03, specialist in palliative medicine) |
| No,  medical school | “Clearly no. Also when it comes to communication with relatives, that is not something you are taught. It is a skill you have to teach yourself. And that is really bitter.” (ID02, specialist in medical oncology and palliative medicine)  “How does someone die? What is it like when someone dies? What does someone die of?” -- You don’t learn that.” (ID10, specialist in medical oncology) |
| No,  postgraduate specialist training (during residency) | “No, not at all. […] In the intensive care unit it was like, ‘Yes, someone is dying, you have to go there now,’ and then I somehow -- I mean, they are deeply sedated with […] a tube in their throat, and I just googled ‘end-of-life care ICU patient’. That is how it went.”(ID10, specialist in medical oncology)  "During residency, I am not really sure, apart from the experience of, let’s say, senior physicians who may have shared their experience or given advice in a particular situation. But in terms of curricular contents, not that I am aware of." (ID21, specialist in hematology/oncology) |
| “Do oncologists have unmet coping support or educational needs regarding patient deaths?” |  |
| No personal unmet needs | “I’ve never had the feeling that I was left alone with the situation or left feeling helpless, not at all. That is why I currently cannot think of anything I would wish for in that regard.” (ID16, resident physician in medical oncology)  “For my area, I think we are set up as optimally as possible.” (ID22, senior/chief physician in medical oncology and palliative care) |
| Timely availability of low-threshold support services | “I think it would be really great if you could turn to something like an anonymous hotline or something like that. That would be awesome.” (ID10, specialist in medical oncology)  “I believe that this [coping support] cannot really be tied to fixed appointments, because ultimately, the situations in which one needs support arise suddenly and unexpectedly. And support is not helpful three weeks later, but what is actually needed is an offer that allows for low-threshold access -- even if only on weekdays or, say, Mondays between 9:00 and 10:00 -- but that one has a contact person with a psychological background who can offer low-threshold counselling.” (ID12, resident physician in medical oncology) |
| Designated spaces to talk about patient death and own emotions | “That […] there is also space for it, if one wants to talk about the fact that one does not feel good with a case. Or that there is also space to say, ‘hey, I cannot do this today, I need to go home now.’” (ID14, specialist in surgery) |
| In medical training |  |
| Practical guidance | “Conversation techniques […] and I think also coping techniques – to be introduced to those would have made sense during medical school, more than it was the case for me.” (ID04, specialist in gynecology)  “If I wanted to try and design this optimally, I believe this is something that should be part of medical school during the final clinical year. That students are taken by the hand and told, ‘imagine you are now the ward physician, could you handle this, and would you know what to do?’” (ID11, senior/chief physician in radiation oncology) |
| Mandatory curricular content | “And I believe it would have been good if, for example, there had been -- in a hospice or palliative care unit -- simply a mandatory module in that area, so that one could have actually seen what it is like there.” (ID07, senior/chief physician in medical oncology and palliative medicine)  “I believe that it actually wouldn’t be bad to make participation in Balint-groups a mandatory part of the specialist training curriculum for residents.” (ID01, senior/chief physician in medical oncology) |
| In the team |  |
| More open communication culture | “I would wish for us to have a healthy approach to dealing with errors, that we would talk through cases, because sometimes there really are things that could be improved.” (ID14, specialist in surgery) |
| Case discussions | “Just, I don’t know, something like once a week, a morning briefing: who died this week -- not just who is being admitted today or what is the surgery schedule --but once, say, on Friday mornings: who passed away this week, how did it go, who was involved, how can we improve things, or something like that.” (ID10, specialist in medical oncology) |
| Supervision groups | “I would also wish for professional supervision on regular wards and especially on intensive care units, where difficult cases can be discussed together as a team.” (ID02, specialist in medical oncology) |
| Team-based farewell rituals | “Once a week, if you just take half an hour in the morning and the head of department is at the front in a big round or something like that […] but that you just say once who has died, that you give death space, that you really give the whole thing a lot of space, I would find that great.” (ID10, specialist in medical oncology) |
| For individuals |  |
| One-on-one conversation offers | “Perhaps it would not be a bad idea if there were one-on-one conversation offers available for every employee.” (ID03, specialist in palliative medicine)  “An official offer that does not take place within the team. Because it certainly has many advantages, but also disadvantages, if you talk to colleagues with whom you continue to work. I think there are things that are not necessarily always easy to address when they affect you very deeply.” (ID18, specialist in palliative medicine) |
| Knowledge and information needs |  |
| Communication training | “Yes, what I would mainly wish for in the workplace is that some kind of training is provided to better deal with palliative patients, how to communicate better with relatives, and just a bit more communication training in general.” (ID13, resident physician in medical oncology) |
| Training programs on death/grief | “And I believe a structured training program [about end-of-life care] is needed. For everyone. And not just for resident physicians, but also for specialists, senior, and chief physicians, because they have not learned it either.” (ID22, specialist in medical oncology and palliative medicine) |
| Dealing with death in different cultures | “Or maybe also -- I find that very important […] -- that there should be a training session on, so to speak, dealing with death in different cultures. What is important for relatives from other cultural backgrounds, where, if you don’t know, you might unintentionally hurt them because you don’t know how to behave appropriately according to their understanding when someone has died. I would find something like that very interesting.” (ID21, specialist in medical oncology) |
| Learning to make better prognoses about life expectancy | “What I would wish for is actually to learn more, also from more experienced colleagues, -- sometimes death is foreseeable […]. I would still say today that I have difficulties assessing how the patient is actually doing at that moment, whether death is truly imminent or if there is still a lot of time. I believe many cannot assess that well, and improving in that regard is something I would wish for.” (ID23, resident physician in medical oncology and surgery) |
