## Supplement I: COREQ Checklist for "Oncologists’ Experiences with Patient Death - a Qualitative Interview Study"

**Supplementary File I:**

**Consolidated criteria for reporting qualitative studies (COREQ): 32-item checklist**

Table A: Consolidated criteria for reporting qualitative studies (COREQ): 32-item checklist ^1^

| **Domain 1: Research team and reflexivity** | | | |
| --- | --- | --- | --- |
| **Personal Characteristics** | | | |
| 1. | Interviewer/facilitator | Which author/s conducted the interview or focus group? | See page 5 |
| 2. | Credentials | What were the researcher’s credentials? E.g. PhD, MD | Medical student |
| 3. | Occupation | What was their occupation at the time of the study? | Medical student, MD candidate; See Supplementary File II |
| 4. | Gender | Was the researcher male or female? | Female |
| 5. | Experience and training | What experience or training did the researcher have? | MG was trained by SW, MR, and IS, who have expertise in qualitative research. |
| **Relationship with participants** | | | |
| 6. | Relationship established | Was a relationship established prior to study commencement? | A relationship for research purposes was established prior to the interview.  See page 5 |
| 7. | Participant knowledge of the interviewer | What did the participants know about the researcher? e.g. personal goals, reasons for doing the research | All reasons were disclaimed.  See Supplementary File II |
| 8. | Interviewer characteristics | What characteristics were reported about the interviewer/facilitator? e.g. Bias, assumptions, reasons and interests in the research topic | MG’s interest in the research topic (doctoral interests) were disclaimed to all participants.  See Supplementary File II |
| **Domain 2: study design** | | | |
| **Theoretical framework** | | | |
| 9. | Methodological orientation and Theory | What methodological orientation was stated to underpin the study? e.g. grounded theory, discourse analysis, ethnography, phenomenology, content analysis | See page 6 |
| **Participant selection** | | | |
| 10. | Sampling | How were participants selected? e.g. purposive, convenience, consecutive, snowball | See page 5 |
| 11. | Method of approach | How were participants approached? e.g. face-to-face, telephone, mail, email | See page 5 |
| 12. | Sample size | How many participants were in the study? | See page 7 and Table 1 |
| 13. | Non-participation | How many people refused to participate or dropped out? Reasons? | See page 5 |
| **Setting** | | | |
| 14. | Setting of data collection | Where was the data collected? e.g. home, clinic, workplace | See page 5 |
| 15. | Presence of non-participants | Was anyone else present besides the participants and researchers? | See page 5 |
| 16. | Description of sample | What are the important characteristics of the sample? e.g. demographic data, date | See page 7 and Table 1 |
| **Data collection** | | | |
| 17. | Interview guide | Were questions, prompts, guides provided by the authors? Was it pilot tested? | See page 5-6 and Supplementary File II |
| 18. | Repeat interviews | Were repeat interviews carried out? If yes, how many? | No |
| 19. | Audio/visual recording | Did the research use audio or visual recording to collect the data? | See page 5 |
| 20. | Field note | Were field notes made during and/or after the interview or focus group? | No |
| 21. | Duration | What was the duration of the interviews or focus group? | See page 5 |
| 22. | Data saturation | Was data saturation discussed? | See page 5 |
| 23. | Transcripts returned | Were transcripts returned to participants for comment and/or correction? | See page 6 |
| **Domain 3: analysis and findings** | | | |
| **Data analysis** | | | |
| 24. | Number of data coders | How many data coders coded the data? | See page 6 |
| 25. | Description of the coding tree | Did authors provide a description of the coding tree? | See Supplementary File III |
| 26. | Derivation of theme | Were themes identified in advance or derived from the data? | See page 6 |
| 27. | Software | What software, if applicable, was used to manage the data? | See page 6 |
| 28. | Participant checking | Did participants provide feedback on the findings? | See page 6 |
| **Reporting** | | | |
| 29. | Quotations presented | Were participant quotations presented to illustrate the themes/findings? Was each quotation identified? e.g. participant number | Each quote was identified with interview number and professional characteristics (medical specialty, clinical role). |
| 30. | Data and findings consistent | Is there consistency between the data presented and the findings? | See pages 7-11 and Supplementary File IV (Tables 2-4) |
| 31. | Clarity of major themes | Were major themes clearly presented in the findings? | See pages 7-11, Supplementary File IV (Tables 2-4) and Figure 1 |
| 32. | Clarity of minor themes | Is there a description of diverse cases or discussion of minor themes? | See pages 7-11 and Supplementary File IV (Tables 2-4) |
