## Supplement II: Interview Guide for "Oncologists’ Experiences with Patient Death - a Qualitative Interview Study"

**Supplementary file II, Interview guide**

| **Verbal consent** | We will now proceed and start our interview. I will switch on the recording devices.  *(MG switches on devices)*  I have now started the audio recording, is that okay with you?  *(Verbal consent?)*  Let’s start the interview then. |
| --- | --- |
| **Subject** | **Probing questions** |
| Introduction  Sharing own experiences (leading over to the experience of patient death in general) | My name is MG, and I am a medical student conducting this study as part of my doctoral thesis in the psycho-oncology research group at the University Medical Center Hamburg-Eppendorf. In our interview today, I would like to ask you about your personal experiences with the death of patients in oncology. It will also be about your coping mechanisms. There will also be questions about how you deal with these experiences. There is no right or wrong here; I am interested in your personal views and experiences.  I would like to ask you to remember a case in which a cancer patient you treated has died. If you like, you can briefly tell me about this case. |
| Experiences with patient death | 1. How did you experience the death you just described, or other patient deaths?    a. How would you describe the feelings and thoughts when you learn or experience that the patient has died / is dying / is going to die soon?    b. How have these feelings developed/changed over time?    c. If you had to give a name to your reactions to patient deaths, how would you call them? 2. Now let’s talk about the topic of death in general:    a. What are your personal beliefs or convictions regarding death?    b. Do you have a specific idea of a "good" death?    c. Have these changed over the course of your experiences with patient death? 3. To what extent does your experience with the death of this patient or of patients in general differ from your experience with private deaths?    a. Are there any similarities between the two experiences?    b. Are there any differences? |
| Impact of experiences with patient death | 4. How did your experiences with the death of patients affect your own life?    a. Were there any professional or private effects?    b. Can you describe these effects to me in more detail? 5. In past cases of deceased patients, how did your surroundings react when you talked about the topic?    a. In a professional context?    b. In a private context?    c. Did you wish for or hope for different reactions? |
| Coping with patient death | 6. What do you do when you learn that a patient has died?    a. Is there anything that you find helpful in these situations? 7. Do you have strategies for coping with patient deaths?    a. Are there any strategies that have proven unhelpful to you? 8. What advice would you give to new/future oncologists who are going to be confronted with the death of cancer patients? |
| Support needs in coping with patient death | 9. Where do you find support for coping with the death of patients when you need it?    a. Are there support offers in your professional environment? 10. Would you wish for (additional) support services? If so, which ones? |
| Educational preparation for coping with patient death | 11. Were you prepared for dealing with the death of patients during the course of your education and professional training?     a. If yes, could you describe what this preparation looked like?     b. If no, how would you have liked that preparation to be?     c. Looking back, would you have wished for a different preparation? Was there something specific missing? 12. Would you currently like further training or educational opportunities on dealing with the death of patients?     a. In what context would you prefer such training? |
| **Ending of the interview**  Open question | 13. Finally, I would like to know if there is anything else you would like to share with me regarding your experiences with the death of patients that I may not have asked you before? |
