## Supplement III: Coding Tree for "Oncologists’ Experiences with Patient Death - a Qualitative Interview Study"

**Supplementary file III, Coding tree**

| Theme | Subtheme  (Level 1) | Subtheme  (Level 2) | Subtheme  (Level 3) | Subtheme  (Level 4) | Description |
| --- | --- | --- | --- | --- | --- |
| **Research question 1: What impacts does patient deathhave on oncologists?** | | | | | |
| Few/ no effects |  |  |  |  | Participant reports no or few noticeable effects from one or more patient deaths; can be coded alongside emotions and/or other effects. |
| Being touched |  |  |  |  | Participant reports that a patient’s death/patient deaths evoke emotions, affecting them emotionally, without a negative connotation. |
| Being distressed |  |  |  |  | Participant reports emotional distress caused by a patient’s death/patient deaths, corresponding to a negative connoted version of “being touched”. |
| Doubts about the legitimacy of own emotions |  |  |  |  | Participant questions the extent to which their own emotional reactions are appropriate. |
| Factors influencing oncologists’ perceived impact of patient deaths |  |  |  |  | “Why do oncologists find some deaths more distressing than others?” |
|  | Related to the mode of exposure to a patient death |  |  |  | Participant reports that the way in which the patient's death is witnessed (e.g., being physically present at the moment of death vs. being retrospectively informed by colleagues) has an impact on the intensity of the experience. |
|  | Confrontation with the limits of medicine |  |  |  | Participant reports that their stress caused by patient deaths is influenced by the experience of being confronted with the limits of current medical possibilities. |
|  | Constraints due to systemic conditions |  |  |  | Participant reports distress due to workplace conditions that make it difficult to provide patient care according to his or her own preferences. |
|  | Patient care not possible according to own moral concepts |  |  |  | Participant reports additional distress if care of dying patients was not possible in accordance with their own moral ideas, e.g., overtreatment of very limited life expectancy. |
|  | Related to clinical setting or medical specialty |  |  |  | Participant reports that the experience of patient deaths is influenced by the treatment setting (e.g., outpatient vs. inpatient) and medical discipline (e.g., intensive care unit vs. palliative medicine settings). |
|  |  | Related to the frequency of experienced patient deaths |  |  | Participant reports a different experience of patient deaths depending on their frequency. |
|  | Related to the nature of death/way of dying |  |  |  | Participant reports increased distress depending on the type of death or the preceding clinical course, e.g., lack of symptom control, unexpected deaths or dramatic courses. |
|  | Related to the physician-patient relationship |  |  |  | Participant reports increased distress depending on the relationship with the patient, e.g., if there has been a long and intensive relationship. |
|  | Related to patient characteristics |  |  |  | Participant reports heightened distress when patients exhibit certain characteristics, e.g., difficulty accepting their impending death or young age. |
|  | Related to characteristics of the patients’ relatives |  |  |  | Participant reports a change in distress depending on characteristics of the patients’ relatives or their own relationship to the relatives. |
|  | Related to characteristics of the oncologist |  |  |  | Participant reports changes in distress caused by patient deaths depending on own characteristics, e.g., professional experience or own similarities with the patient. |
| Emotional impact |  |  |  |  |  |
|  | Satisfaction |  |  |  | Participant reports feelings of satisfaction, e.g., with their own work, the quality of provided care. |
|  | Relief |  |  |  | Participant reports a feeling of relief, e.g., because the suffering of the deceased patient has ended as a result of the death. |
|  | Gratitude/appreciation |  |  |  | Participant reports feeling grateful, e.g., for the time spent together or the trust of the patient/relatives. |
|  | Existential sorrow (*Weltschmerz)* |  |  |  | Participant reports feeling disheartened, e.g., in light of life’s unfairness. |
|  | Overwhelm/helplessness |  |  |  | Participant reports feeling helpless, overwhelmed, or left alone: “resources do not match tasks”. |
|  | Sadness |  |  |  | Participant reports feeling sad about a patient death. |
|  | Shock |  |  |  | Participant reports feeling surprised, taken off-guard or shocked by a patient death. |
|  | Anger |  |  |  | Participant reports feelings of anger, e.g., towards colleagues or the circumstances under which care had to be provided. |
|  | Guilt |  |  |  | Participant reports feelings of guilt relating to the death or inadequacies in the treatment of the patient. |
|  | Grief |  |  |  | Participant reports feelings of grief over the death of a patient.  . |
|  | Compassion |  |  |  | Participant reports feeling compassion. |
|  |  | For patients |  |  | Participant reports compassion for patients. |
|  |  | For relatives |  |  | Participant reports compassion for relatives. |
|  | Fear of one's own finiteness |  |  |  | Participant reports fear of their own finiteness being evoked by (frequent) confrontation with the patient deaths. |
| Cognitive impact |  |  |  |  |  |
|  | Long-term vivid memories of deceased patients |  |  |  | Participant reports long-term detailed memories of deceased patients. |
|  | Recurring thoughts of deceased patients |  |  |  | Participant reports recurring or persistent thoughts of deceased patients. |
|  | Sense of responsibility |  |  |  | Participant reports an increased awareness of medical responsibility in connection with patient deaths. |
|  | Conscious experience of life’s finality |  |  |  | Participant reports a conscious realization of the finality of death through experiences with patient deaths. |
| Professional impact |  |  |  |  |  |
|  | Routine in dealing with the patient death |  |  |  | Participant reports having developed a routine in dealing with the death of patients with frequent confrontation leading to a certain habituation, which helps to maintain composure. |
|  | Proactive communication with patients about death and dying |  |  |  | Participant reports having decided to communicate more proactively with dying patients about death and placing more emphasis on preparing death with patients. |
|  | Decision to pursue additional training/further education |  |  |  | Participant reports that, as a consequence of experiences with patient deaths, they decided to pursue additional qualifications, such as in palliative medicine. |
|  | Reduction of work in clinical patient care |  |  |  | Participant reports that, as a consequence of experiences with patient deaths, they have partially or completely withdrawn from clinical work. |
|  | Self-doubts about one’s competence |  |  |  | Participant reports doubting their own professional competence in caring for patients as an oncologist; feelings of inadequacy. |
|  | Decreased emotional resonance/empathy towards patients |  |  |  | Participant reports feeling less able to empathize with patients due to the confrontation with patient deaths. |
| Personal life impacts |  |  |  |  |  |
|  | Greater appreciation for one’s own life (perspective taking) |  |  |  | Participant reports that engaging with patient death has helped them to value their own life and health (or that of relatives) more, and has supported them in setting personal priorities and making life decisions that feel right to them (with the awareness that life is finite). |
|  | Changes in beliefs related to death |  |  |  |  |
|  |  | Dissolution/loss of previous beliefs |  |  | Participant reports loss or dissolution of (religious) beliefs; does not have to be subjectively seen as negative. |
|  |  | Consolidation/clarification of one’s own beliefs |  |  | Participant reports changes in beliefs and evaluates these changes clearly as positive. |
|  | Serenity in dealing with death and dying |  |  |  | Participant reports that frequent confrontation with death and dying in their professional life has led to greater calm in dealing with the death in personal life. |
|  | Active engagement with death in one’s private life |  |  |  | Participant reports that, due to confrontation with patient deaths, they have begun to prepare more actively for their own or their relatives’ death, e.g., through conversations or practical measures. |
|  | Difficulties in separating personal and professional life |  |  |  | Participant reports difficulties in maintaining boundaries between professional and private life. |
| Distinction between experiences with death in professional and personal contexts |  |  |  |  |  |
|  | Difference in the duration of emotional impact |  |  |  | Participant reports that emotional responses to death in professional contexts tend to be shorter in duration. |
|  | Difference in intensity |  |  |  |  |
|  |  | Due to greater emotional regulation in response to patient death |  |  | Participant reports that emotional self-regulation is easier in response to patient death; this may be due to the greater demands placed on one’s own regulatory capacity (professionalism). |
|  |  | Due to less close personal connection with patients |  |  | Participant reports differences in emotional intensity, attributed to a less close bond with patients compared to people from their private life. |
|  |  | Professional role affected vs. entire self affected |  |  | Participant reports that personal losses affect the entire self emotionally whereas the emotional impact of patient death is “limited” to the professional role. |
|  | Commonality: Experience of grief/sadness |  |  |  | Participant reports experiencing sadness as a shared element in both private and professional encounters with death. |
| **Research question 2: How do oncologists cope with patient death?** | | | | | |
| Coping strategies |  |  |  |  |  |
|  | Intrapersonal strategies |  |  |  | “no other person necessary” |
|  |  | Activities |  |  |  |
|  |  |  | Professional reflection on patient death |  | Participant reports that, after a patient death, they like to reflect on the case from a medical perspective and retrospectively evaluate treatment decisions, also in order to potentially improve future patient care. |
|  |  |  | Closing information gaps about patient death |  | Participants report that, in cases of missing information about the exact course of death, they seek clarification by gathering information. |
|  |  |  | Continuing work tasks |  | Participant reports quickly returning to work routine. |
|  |  |  | Lighting a candle |  | Participant reports lighting a candle as a coping strategy. |
|  |  |  | Visiting a spiritual space |  | Participant reports visiting a spiritual place to process the death of a patient, e.g., a cemetery or a chapel. |
|  |  |  | Physical activity |  | Participant reports engaging in physical activity to cope with patient death, e.g., walking or sports. |
|  |  | Adapting one’s attitude |  |  |  |
|  |  |  | Consciously allowing emotional connection and vulnerability with patients |  | Participant reports consciously allowing close relationships with patients and accepting their own vulnerability as part of this. |
|  |  |  | Applying own beliefs about life and death |  | Participant reports relying on established personal, including religious, beliefs to cope with patient deaths. |
|  |  |  | Acceptance |  | Participant reports that acceptance helps them deal with patient deaths. |
|  |  | Finding the right balance |  |  |  |
|  |  |  | Letting time pass |  | Participant reports that consciously letting time pass helps in coping with patient death. |
|  |  |  | Pausing/taking a moment |  | Participant reports taking short pauses or moments for oneself to cope with patient death. |
|  |  |  | Seeking distraction or balance |  | Participant reports using distraction or engaging in leisure activities as a strategy to balance the emotional impact of patient death. |
|  |  |  | Seeking physical distance from work |  | Participant reports that leaving the workplace and “the way home” serve as a coping strategy; physical distance between work and private life is perceived as helpful. |
|  |  |  | Maintaining a professional distance |  | Participant reports that maintaining professional distance before and after patient death helps them cope. The extent of this distance depends on subjective perception and may be seen as either functional or dysfunctional. |
|  |  |  | Clear separation between personal and professional life |  | Participant reports maintaining clear boundaries between work and private life as a coping strategy. |
|  |  |  | Humor |  | Participant reports using (dark) humor as a coping strategy. |
|  | Interpersonal strategies |  |  |  | Presence of another person is necessary in some way. |
|  |  | Professional support |  |  | Participant reports using formalized support services, such as supervision, peer consultation groups, or psychological/pastoral counseling to cope with patient death. |
|  |  |  | Participation in group/team formats |  | Participant reports making use of group-based support services, such as Balint-groups. |
|  |  |  | Support from chaplains |  | Participant reports receiving support from chaplains. |
|  |  |  | Support from psychologists |  | Participant reports receiving support from psychologists. |
|  |  | Informal social support |  |  | Participant describes informal social support as a coping resource. |
|  |  |  | Support from colleagues |  | Participant reports receiving informal social support from colleagues of different professions. |
|  |  |  |  | From physicians | Participant reports informal support among medical colleagues as a coping strategy. |
|  |  |  |  | From nursing staff | Participant reports informal support from nursing colleagues as a coping resource. |
|  |  |  |  | From psychological colleagues | Participant reports receiving informal support from psychological colleagues as a coping strategy. |
|  |  |  |  | From other professional groups | Participant reports receiving informal support from other professional groups (e.g., social workers, art therapists) as a coping strategy. |
|  |  |  | Support from private social circle |  | Participant reports social support from their partner, friends, or family as a coping resource. |
|  |  | Expression of emotions |  |  | Participant reports that expressing or sharing their own emotions helps them cope with patient death. |
|  |  | Saying goodbye to the body |  |  | Participant reports that (re)visiting the body of the deceased person helps them cope. |
|  |  | Initiating contact with patients’ relatives |  |  | Participant reports contacting relatives of deceased patients, e.g., to offer condolences or to express availability for further questions or conversations. |
|  | Rituals |  |  |  | Any coping strategy carried out in a ritualized form, usually repeated over time. |
|  |  | Googling obituary notices |  |  | Participant reports regularly searching online for obituary notices after patient deaths. |
|  |  | Acknowledgement/honoring the deceased patient |  |  | Participant states that it is important to commemorate in some form, as part of a personally chosen ritual. |
|  |  | Team-based/institutional rituals |  |  | Participant reports taking part in mourning rituals organized by the team or institution, e.g., death rounds or memorial services for patients. |
|  |  | Praying |  |  | Participant reports praying as a ritualized coping strategy. |
| Coping barriers |  |  |  |  | All aspects impeding oncologists’ successful coping with patient deaths. |
|  | Lack of professional support services |  |  |  | Participant reports that no support services are available in their professional environment in the event of patient deaths. |
|  | Taboo around death |  |  |  | Participant reports that a perceived taboo surrounding death acts as a barrier to dealing with patient deaths. |
|  | Limited acceptance of existing professional support services |  |  |  | Participant reports that they or colleagues do not sufficiently make use of available support services |
|  |  | - Lack of awareness of support services and access pathways |  |  | Participant reports that support services may be available through the employer, but that they or colleagues are unsure how to access them or who to contact. |
|  | Sharing experiences perceived as unhelpful |  |  |  | Participant reports experiences with unhelpful conversations about patient deaths, e.g., due to a lack of empathy, understanding, or emotional capacity of the conversation partner. |
|  |  | With people from private life |  |  | Social exchange with people from participants’ private circle perceived as unhelpful. |
|  |  | With colleagues |  |  | Social exchange with colleagues or superiors perceived as unhelpful. |
|  |  |  | In supervision |  | Supervision formats perceived as unhelpful. |
|  | No expression of emotions |  |  |  | Participant reports that it is not possible to express their emotional responses to patient deaths, e.g., because conversation partners are unavailable. |
|  | Time pressure |  |  |  | Participant reports feeling they do not have enough time to feel and express their emotions or to say goodbye, e.g., due to a hectic schedule or the urgency of other tasks. |
|  | Pressure to meet expectations/perfectionism in dealing with patient death |  |  |  | Participant reports feeling pressured by their own or others’ expectations that there is a "right" way to deal with patient deaths. |
|  | Suppressing one’s own emotions |  |  |  | Participant reports not wanting to feel their own emotional responses to patient death, e.g., by avoiding situations that might trigger them or by cognitively suppressing feelings. |
|  | Dishonest communication with patients about prognosis |  |  |  | Participant reports inadequate communication by oncologists with patients by giving unrealistic hope or avoiding honest discussion of prognosis. |
|  | Blame dynamics |  |  |  | Participant reports that blame dynamics among colleagues and superiors act as a barrier to helpful ways of dealing with patient deaths. |
|  | Perceived threat of legal consequences from treatment decisions |  |  |  | Participant reports that fear of legal consequences related to treatment decisions, as well as advice aimed at avoiding such consequences, can add additional emotional burden. |
| **Research question 3: Were oncologists educationally prepared to cope with patient death? If so, where?** | | | | | |
| YES,  Medical school |  |  |  |  | Participant reports having received some form of preparation for dealing with patient death during university studies (medical school). |
|  | Pre-final year |  |  |  | Participant reports having received preparatory content on patient death as part of the preclinical of medical school. |
|  | Final clinical year |  |  |  | Participant reports having been prepared for patient death during the final clinical year (“Practical Year” in German medical education) of medical school. |
| YES,  Postgraduate specialist training (during residency) |  |  |  |  | Participant reports having received instructions on dealing with patient death during postgraduate specialist training. |
| NO,  Medical school |  |  |  |  | Participant reports not having been prepared for dealing with patient death during medical school, or having felt insufficiently prepared. |
| NO,  Postgraduate specialist training (during residency) |  |  |  |  | Participant reports not having been prepared (sufficiently) for dealing with patient death during postgraduate specialist training. |
| **Research question 4: Do oncologists have unmet coping support or educational needs regarding patient death?** | | | | | |
| NO personal unmet needs |  |  |  |  | Participant reports having no current unmet needs related to patient death. |
| Timely availability of low-threshold support services |  |  |  |  | Participant reports that it would be helpful to receive timely support after patient death, e.g., in the form of a psycho-oncological or spiritual care hotline. |
| Designated spaces to talk about patient death and own emotions |  |  |  |  | Participant reports wishing for a designated space to talk about one’s experiences and emotions related to patient death. |
| In medical training |  |  |  |  |  |
|  | Practical guidance |  |  |  | Participant reports wishing for more practical guidance during medical training, e.g., in the final clinical year on how to deal with patient death. |
|  | Mandatory curricular content |  |  |  | Participant reports wishing for mandatory modules on dealing with patient death at different stages of medical education. |
| In the team |  |  |  |  |  |
|  | More open communication culture |  |  |  | Participant reports wishing for more open communication within the team and with supervisors about patient deaths and about opportunities to improve care (e.g., through error management). |
|  | Case discussions |  |  |  | Participant wishes for case discussions on deceased patients, including to critically reflect on their own practice. |
|  | Supervision groups |  |  |  | Participant wishes for (more frequent) team supervision. |
|  | Team-based farewell rituals |  |  |  | Participant expresses a desire to participate in team-based rituals to say goodbye to deceased patients. |
| For individuals |  |  |  |  |  |
|  | One-on-one conversation offers |  |  |  | Participant reports needing more structured opportunities for individual conversations about their own grief in connection with patient death. |
| Knowledge and information needs |  |  |  |  |  |
|  | Communication training |  |  |  | Participant expresses interest in support offers to improve communication with patients and relatives about death. |
|  | Training programs on death/grief |  |  |  | Participant wishes for workshops on death or grief. |
|  | Dealing with death in different cultures |  |  |  | Participant reports wishing for training on dealing with death in different cultural contexts to better address the needs of diverse patients and relatives. |
|  | Learning to make better prognoses about life expectancy |  |  |  | Participant reports wishing to improve their skills and knowledge regarding prognosis and estimating remaining life expectancy in patients. |
